# Development and validation of an electronic health records-based opioid use disorder algorithm by expert clinical adjudication

**DOI:** 10.1101/2021.09.23.21264021

**Authors:** Shabbar I. Ranapurwala, Ishrat Z. Alam, Brian W. Pence, Timothy S. Carey, Sean Christensen, Marshall Clark, Paul R. Chelminski, Li-Tzy Wu, Lawrence H. Greenblatt, Jeffrey E. Korte, Mark Wolfson, Heather E. Douglas, Lynn A. Bowlby, Michael Capata, Stephen W. Marshall

## Abstract

**Background:** In the US, over 200 lives are lost from opioid overdoses each day. Accurate and prompt diagnosis of opioid use disorders (OUD) may contribute substantially to prevention of overdose deaths. However, OUD research is limited, the specificity and sensitivity of OUD ICD codes are unknown, and the ICD codes are known to underestimate OUD prevalence. We developed and validated algorithms to identify OUD from EHR data and examine validity of ICD-based definitions for OUD.

**Methods:** Through multiple iterations, we developed EHR-based algorithms to identify OUD. These algorithms and ICD-based OUD definition were validated against a total of 169 independent gold standard EHR chart reviews conducted by an expert adjudication panel of eight pain and addiction medicine clinical experts across four large healthcare systems. The experts relied on clinical judgement and current Diagnostic and Statistical Manual of Mental Disorders-5 criteria for making OUD diagnoses.

**Results:** Of the 169 EHR charts, 81 (48%) were reviewed by more than one expert and exhibited 85% agreement between the reviewing experts. The OUD ICD codes alone had 10% sensitivity and 99% specificity, underscoring the strong potential for OUD underestimation in studies depending on ICD codes alone. In comparison, after four iterations, the algorithms identified OUD with a 23% sensitivity and 98% specificity.

**Conclusions and Relevance:** This is the first study to evaluate the validity of OUD ICD codes and develop validated EHR-based algorithms to address OUD underestimation. This work has the potential to inform future research on early intervention and prevention of OUD.

## Introduction

Each day more than 200 individuals die from opioid overdoses in the United States (US).^1,2^ The overdose death estimates represent only the tip of the opioid epidemic iceberg, with >2 million Americans suffering from opioid use disorders (OUD), a term which encompasses addiction, abuse, and dependence. Another 10 million or more Americans are misusing opioids and are at risk of developing OUD.^3^ Even as most opioid research to date has focused on preventing overdose deaths,^4–7^ opioid overdose deaths from prescription and illicit opioids have increased.^1,2^ Continued progress in combatting the opioid epidemic requires a shift towards earlier intervention to prevent and treat opioid use disorder.^8^

The limited research on OUD prevention derives from difficulties in reliably identifying OUD in large healthcare databases.^9,10^ ICD codes for OUD exist but are likely under-utilized because OUD can be challenging to identify clinically^11^ and the stigma of an OUD diagnosis may negatively affect patients’ health insurance, employment, and healthcare. As a result, ICD codes may have low sensitivity in identifying OUD and contribute to underestimation and under-treatment for OUD.^12,13^ Clinical review of patients’ medical records can allow more accurate OUD identification rather than relying on ICD codes alone, but it is a time-intensive strategy, beyond the resources for most research projects and infeasible in large healthcare databases.^12^

The early and accurate identification of patients with OUD is critical in linking patients to treatment to prevent overdose deaths, develop OUD prevention strategies by examining its predictors, and reduce suffering and loss of productivity due to medical comorbidities.^9–13^ Using electronic health record (EHR) data from four large healthcare systems in two US states, we iteratively developed and validated algorithms to identify OUD from EHR data. We used expert-adjudicated OUD diagnosis as a gold standard. We compared sensitivity and specificity for identifying OUD using our algorithms and using of ICD codes alone.

## Methods

We conducted a validation study in four large academic healthcare systems including the University of North Carolina at Chapel Hill, Duke University, Wake Forest Baptist Health, and Medical University of South Carolina. We used structured EHR data from 2014-2017 and variables available in the PCORnet common data model^14^ to increase the generalizability of the algorithm. The variables included age, gender, race, ethnicity, ICD 9/10 diagnosis codes, encounter information, medications, procedures, and referral codes.

### Gold standard

To develop and validate the algorithms to identify OUD and examine the validity of the ICD codes alone, we established a gold standard expert adjudication panel comprising two experts at each institution (8 total). The experts included psychiatrists with addiction training, pain medicine specialists, general internal medicine physicians with experience in treating OUD, and substance use disorder treatment specialists. All experts reviewed and applied clinical judgement to the 11 criteria for OUD from the Diagnostic and Statistical Manual of Mental Disorders-5 (DSM-5); meeting any two of the 11 criteria is considered sufficient for a diagnosis of OUD (APA, 2013) – experts used clinical judgement to ascertain whether the patients met these criteria or not.^15^

### Algorithm building and validation

**Figure 1.** shows a schematic of the methods involved in developing the algorithm to identify OUD. We first identified a cohort of all new patients, defined as a patient with no medical encounters in the 6 months prior to the first observed encounter (index encounter) between 2014 and 2017. We then identified new patients who received at least two opioid prescriptions during any moving 6-month period subsequent to the index encounter. This represented 10.5% of all new patients seen at all four institutions.

**Figure 1.**
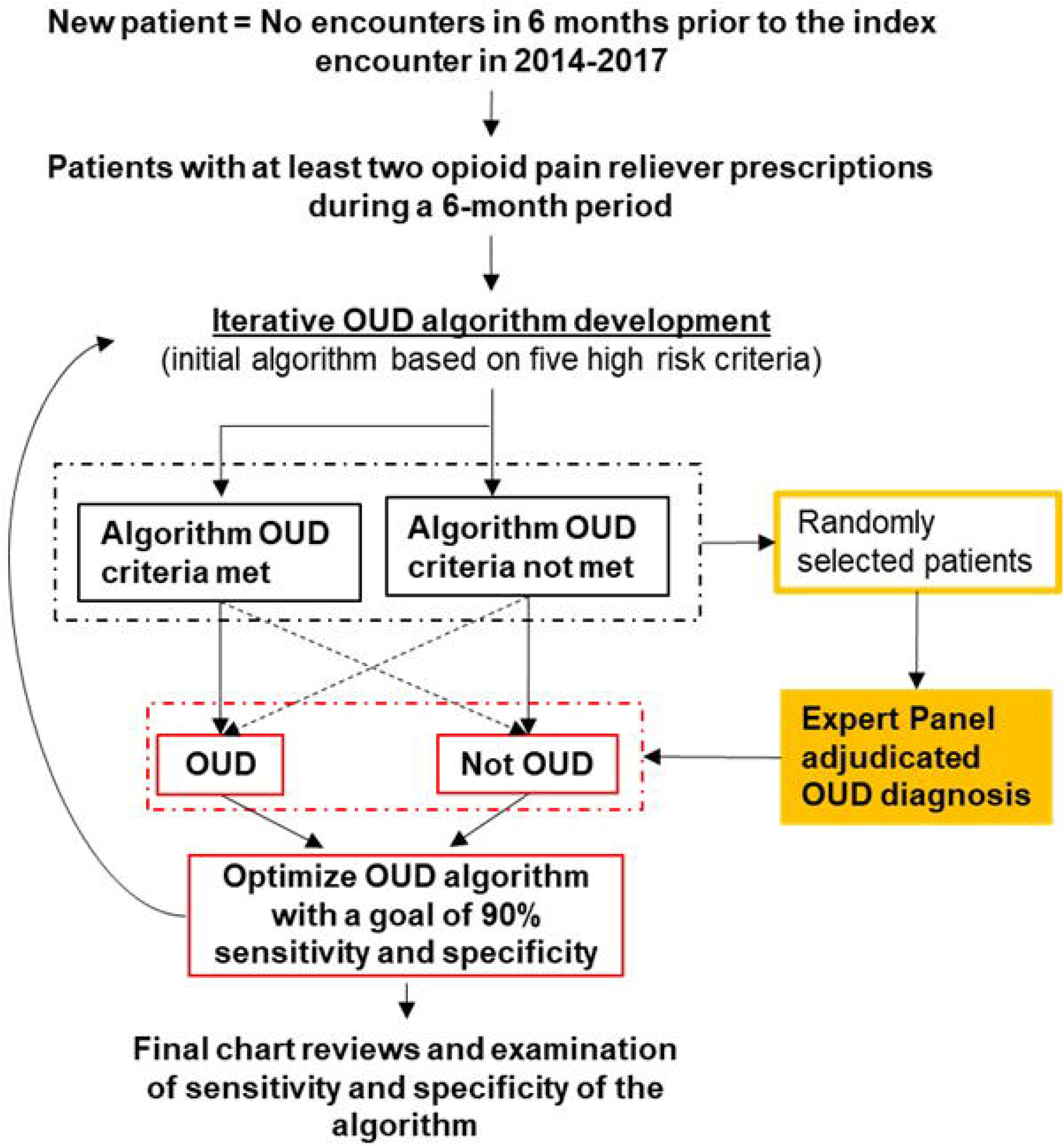
Patient selection and algorithm development.

We used this cohort of new patients with at least two opioid prescriptions in a 6-month period for developing algorithms to determine OUD status. Our initial (stage-1) algorithm included five criteria (Table 1), intentionally designed to be highly sensitive rather than specific. Patients who met at least one of these criteria during 2014-2017 were classified as probable OUD patients, while patients who met none of the criteria were classified as probable non-OUD patients.

**Table 1.** Opioid use disorder algorithm iterations, definitions, and performance compared to expert panel adjudication.

| Iteration | # Charts reviewed | Criteria | PPV<br>(True + / Algorithm +) | NPV<br>(True - / Algorithm -) |
| --- | --- | --- | --- | --- |
| 1 | 40 | Any of: A, B, C, D, E | 50% (10/20) | 90% (18/20) |
|  | 40 | A only (ICD codes) | 71% (2/7) | 79% (26/33) |
| 2* | 38 | Any of: A, B, C, D, E | 32% (6/19) | 89% (17/19) |
|  | 38 | A only (ICD codes) | 50% (4/8) | 87% (26/30) |
| 3 | 46 | Any of: A, D, F, G, H | 71% (15/21) | 88% (22/25) |
|  | 46 | A only (ICD codes) | 100% (12/12) | 82% (28/34) |
|  | 46 | Iteration 3 minus F | 83% (15/18) | 89% (25/28) |
| 4** | 45 | Any of: A, D, F, G, I | 65% (15/23) | 95% (21/22) |
|  | 45 | A only (ICD codes) | 85% (11/13) | 84% (27/32) |
|  | 45 | Iteration 4 minus F | 70% (14/20) | 92% (23/25) |

A. ICD9/10 codes for abuse/ dependence
B. transition to buprenorphine, methadone, and naltrexone that were prescribed to treat OUD or co-prescription of Naloxone to prevent overdose
C. $\geq 50$ MME of opioids for $\geq 180$ days
D. procedure codes for Medication Assisted Treatment for OUD
E. $\geq 1$ overdose event during the study period
F. $\geq 90$ MME of opioids for $\geq 180$ days
G. 'B'+ 'E'
H. 3+ ED visits with opioid RX in 30-day window
I. 'H,' excluding ED visits where surgery was performed up to two weeks before visit

We randomly selected 10 stage-1 patients, 5 probable OUD and 5 probable non-OUD, from each institution (40 total). The expert panel reviewed full clinical details and triage notes from these patients without the stage-1 classification results and adjudicated the cases. Each reviewer was allotted eight of the 10 site-specific cases such that six cases were reviewed by two reviewers at each site.

After each case was reviewed, the experts provided a decision of OUD diagnosis or no OUD diagnosis and reported the factors that met the DSM-5 OUD criteria or other factors they used to decide that the patient did or did not have an OUD.

We then compared expert-adjudicated OUD diagnoses with our stage-1 algorithm results and calculated stage-1 sensitivity and specificity. To improve the algorithm’s performance, we refined the stage-1 algorithm by incorporating the factors experts used for their decision making, particularly in discordant cases (algorithm positive/expert negative or vice versa). The resulting stage-2 algorithm was then utilized to repeat the whole process, including identifying probable OUD cases in the original cohort of patients, randomly selecting a new set of 10 patients from each site for expert adjudication, and examining validity of the algorithm. We repeated this process for four iterations. Details about the iterations and coding can be accessed here: https://github.com/ShabbarIR/OUD-algorithms-development-and-validation.

### Statistical analysis

We report positive and negative predictive values (PPV and NPV) of each stage of algorithm iteration, as well as for using ICD codes alone for OUD based just on the cases reviewed at that stage. Following this, we used the total sample of all patients with adjudicated OUD status from any of the four algorithm iterations as the full sample to evaluate the final performance of stage-3 and stage-4 algorithms as well as for ICD 9/10 codes alone. Since the expert adjudication was conducted in a sample of the charts, we used inverse probability of sampling proportion (IPSP) weights to adjust the sensitivities and specificities of the algorithms.^16^ We present weighted sensitivity, specificity, PPV, and NPV with exact 95% confidence intervals (CI) and likelihood ratios (LR+/LR−).^17,18^

We estimated adjusted OUD prevalence using the sensitivity and specificity estimates to quantify the OUD prevalence underestimation from our algorithms and the ICD codes (Table 3), using the following formula:^19^

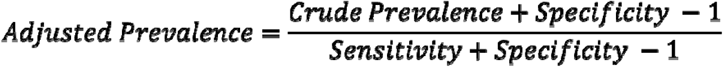

We used Cohen’s kappa^20^ to examine agreement between the reviewers. One reviewer dropped out during stage-2 leaving only one reviewer at one of the sites. At that site Cohen’s kappa was calculated using the stage-1 results only.

To ensure consistency throughout the study procedures, EHR extraction, programming, and analysis steps were developed at one institution, and then the SQL/SAS codes were shared with the other three institutions.

## Results

A total of 169 charts were reviewed by the expert adjudication panel to validate the algorithms and the ICD codes. During stage-2, one reviewer at a site dropped out, so only eight charts were reviewed there; and during stage-3 and stage-4 iterations, a programming error at one site necessitated the review of an additional 11 charts during those two iterations. The Cohen’s kappa for agreement between the site experts was 0.65 (95% confidence interval: 0.46-0.83). The average observed agreement between reviewers was 85.2% with a minimum of 83.3%.

The stage-3 unweighted PPV (71%) and NPV (88%) along with increased PPV and NPV suggest that it is the most balanced algorithm (Table 1) and stage-4 algorithm, which excludes cancer patients, has the highest unweighted NPV (95%). The ICD-based OUD definition produced a 100% unweighted PPV (at stage-3) and 82% unweighted NPV suggesting underestimation of OUD prevalence.

When we used IPSP weights to adjust for the stratified sampling, we found that the sensitivity of both stage-3 (23.5%) and stage-4 (16.7%) algorithms and the ICD-based OUD definition (10.3%) was very low (Table 2). The specificity of both the algorithms and the ICD-based OUD definition were 98.3%, 98.5%, and 99.5%, respectively. Thus, compared to ICD-codes, a small decrease in specificity increased the sensitivity of the OUD algorithms by a factor of 1.6 to 2.3 times.

**Table 2.** Validity of stage 3 and 4 iterations and ICD9/10 codes for OUD using all charts and inverse probability of sampling proportion weights and exact 95% confidence intervals.

| Iteration | Criteria | Sensitivity<sup>\$</sup><br>(95%CI) | Specificity<sup>\$</sup><br>(95% CI) | PPV<sup>\$</sup><br>(95% CI) | NPV<sup>\$</sup><br>(95% CI) | Likelihood<br>ratio+ | Likelihood<br>ratio- |
| --- | --- | --- | --- | --- | --- | --- | --- |
| 3 | Any of: A, D, F, G, H | 23.54%<br>(23.33-23.74) | 98.29%<br>(98.27-98.32) | 61.35%<br>(60.98-61.72) | 91.79%<br>(91.75-91.83) | 13.77 | 0.78 |
|  | A only (ICD codes) | 10.41%<br>(10.28-10.53) | 99.48%<br>(99.47-99.50) | 79.61%<br>(79.15-80.06) | 85.11%<br>(85.05-85.17) | 20.02 | 0.90 |
|  | Iteration 3 minus F | 17.91%<br>(17.73-18.09) | 98.72%<br>(98.70-98.73) | 61.62%<br>(61.19-62.05) | 91.27%<br>(91.23-91.32) | 13.99 | 0.83 |
| 4* | Any of: A, D, F, G, I | 16.67%<br>(16.49-16.84) | 98.48%<br>(98.46-98.50) | 57.54%<br>(57.11-57.96) | 90.54%<br>(90.49-90.58) | 10.97 | 0.85 |
|  | A only (ICD codes) | 9.85%<br>(9.72-9.97) | 99.61%<br>(99.60-99.62) | 82.17%<br>(81.69-82.64) | 85.88%<br>(85.82-85.94) | 25.25 | 0.91 |
|  | Iteration 4 minus F | 13.61%<br>(13.46-13.77) | 98.87%<br>(98.85-98.89) | 59.78%<br>(59.31-60.25) | 90.26%<br>(90.22-90.31) | 12.04 | 0.87 |

A. ICD9/10 codes for abuse/ dependence
B. transition to buprenorphine, methadone, and naltrexone that were prescribed to treat OUD or co-prescription of Naloxone to prevent overdose
C. ≥50 MME of opioids for ≥180 days
D. procedure codes for Medication Assisted Treatment for OUD
E. ≥1 overdose event during the study period
F. ≥90 MME of opioids for ≥180 days
G. 'B'+E'
H. 3+ ED visits with opioid RX in 30-day window
I. 'H,' excluding ED visits where surgery was performed up to two weeks before visit

The weighted PPV and NPV estimates suggest (Table 2), that even at seemingly high values, the ICD codes and our algorithms can have low sensitivity and greatly underestimate the true prevalence of disease (Table 3). Based on the low weighted sensitivity of the algorithms, the overall prevalence of OUD was estimated to be between 14-15% (Table 3) among new patients who received at least two opioid prescriptions during a 6-month period subsequent to their index encounter between 2014-2017. The crude prevalence estimate from the stage-3 algorithm was found to be the least underestimated, and yet it underestimated OUD prevalence by over 5 times (Table 3).

**Table 3:**
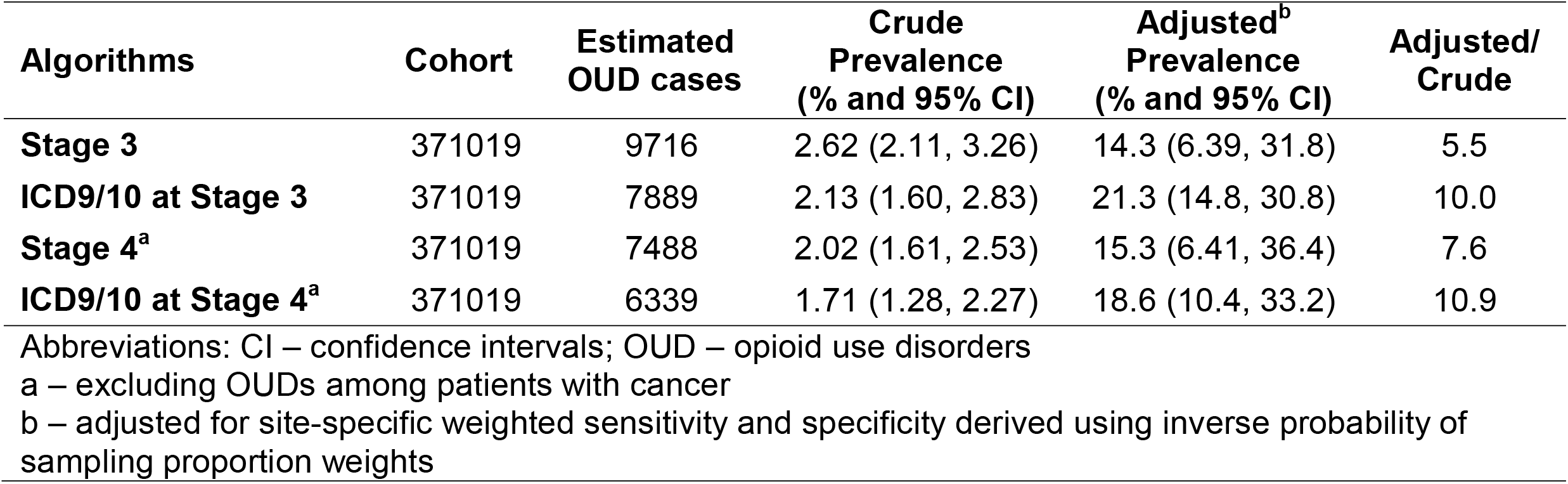
Crude and adjusted OUD prevalence estimates.

## Discussion

OUD has a significant impact on patients, families, and healthcare delivery systems.^21^ Improved OUD case identification may help to intervene early in the opioid use cycle, thereby facilitating targeted prevention of OUD and overdoses. This is the first study to define and validate EHR-based OUD algorithms using expert clinical adjudication as a gold standard, and the first study to estimate the validity of ICD-based OUD definitions. Prior studies have found ∼58%-62% PPV for the ICD-based OUD definition, but none so far have examined sensitivity of these codes.^9,10^ Our study underscores how seemingly reasonable estimates of PPV may be associated with very low sensitivity and underestimate the scale of a problem. With little loss of specificity, our algorithms improve upon the low sensitivity of ICD-based OUD definition and provide the foundation for future research refinements to build more sensitive algorithms. Future algorithm refinements could help healthcare delivery systems identify patients with probable OUD who could benefit from further evaluation and linkage to appropriate care.

The use of the highly specific ICD-based OUD definition yields fewer false positive diagnoses; however, its low sensitivity misses the majority of patients with OUD (high false negative rates). This is problematic in devising robust clinical and population-based responses to stem the opioid epidemic. Implementing an algorithm with greater sensitivity optimizes opportunities for treating a condition with low prevalence such as OUD.^12,13^

This study is the first to allow for stratification of the algorithm to include or exclude cancer patients who are being treated with opioids. Management of pain for cancer patients is complex, often involving collaborative care models to prescribe opioids for pain management in hospice settings. The ability to exclude cancer patients and evaluate OUD among non-cancer patients is therefore invaluable.^4,5^

Our adjusted OUD prevalence estimates are much higher than estimates from the National Survey on Drug Use and Health (NSDUH) of self-reported OUD and opioid misuse prevalence in the general population.^3^ However, note that our estimates are among people who received at least two opioid prescriptions in a six-month period (10.5% of all new patients). Adjusting for this population selection, our estimates of OUD prevalence in the general population would be slightly lower than NSDUH’s estimates,^3^ and perhaps more clinically relevant than the NSDUH estimates.

### Limitations

Our algorithms may not be sensitive to people who primarily use illicit opioids, because these individuals may not consistently encounter the healthcare system. Yet, prescription opioid misuse is often a gateway to using illicit opioids,^6–8^ and early identification of prescription-related OUD may prevent new illicit opioid use. In addition, some of the criteria used in our algorithms may change over time due to the dynamic nature of the opioid epidemic. However, incorporating such algorithms in healthcare delivery systems can allow future improvements through machine learning and artificial intelligence methods. Finally, our algorithms are validated in four healthcare systems. Though they represent diverse patient populations, further testing in healthcare settings across the US will improve generalizability. The algorithms are primed for such testing since we used the PCORnet common data model to develop them.

## Conclusions

Using expert clinical adjudication as a gold standard, we underscore the substantial underestimation of OUD prevalence when using ICD-based OUD definitions in EHR data and present the validity of more refined EHR-based OUD algorithms. These estimates and algorithms will allow adjustment of findings from previous studies using quantitative bias correction methods^17,22^ and facilitate new research focused on early intervention on OUD prevention and treatment to respond to the ongoing opioid epidemic.^4–7^

## Data Availability

The data from the study are not directly available as these contain private health information obtained from electronic medical records. Aggregate level deidentified data can be made available by emailing the corresponding author. All code and methods used are available online at: https://github.com/ShabbarIR/OUD-algorithms-development-and-validation

